# Real-time Computer Vision Assisted Navigation for Endoscopic Pituitary Surgery: Iterative Development and Comparative Preclinical Evaluation

**DOI:** 10.64898/2026.06.02.26354760

**Authors:** Danyal Z Khan, Zhehua Mao, George Hudson, Anjana Wijekoon, Danny Chen, Anouk Borg, Neil Dorward, Ann Blandford, Matt Clarkson, Peter McCulloch, Sophia Bano, Danail Stoyanov, Hani J Marcus.

**Author notes:** joint first (Danyal Z Khan, Zhehua Mao).

## Abstract

**Background:** Endoscopic pituitary surgery involves navigating high-stakes anatomy where complications, such as carotid artery injury, cause devastating morbidity. While computer vision AI offers potential for real-time anatomical recognition to mitigate these risks, successful translation requires rigorous human-factors and performance evaluation. We present the iterative development and preclinical evaluation of a surgeon-controlled, real-time AI-assisted navigation system.

**Methods:** Guided by IDEAL Stage 0 and DECIDE-AI frameworks, the study was conducted in two phases. Phase 1 was an exploratory study where surgeons used the system during high-fidelity simulated surgery and provided feedback via “Think Aloud” protocols and surveys. Following prototype iteration, a Phase 2 randomized crossover comparative trial was conducted with 19 neurosurgeons (15 trainees, 4 experts) performing high-fidelity simulated tumour resections with and without AI assistance, separated by a minimum 2-week washout. The primary outcome was surgical technical performance (OSATS). Workload, educational value, usability, trust, and implementation outcomes were also assessed.

**Results:** Phase 1 informed hardware, model, and interface refinements, including optimized pedal-controlled overlays and prediction confidence metrics. In the comparative trial, AI assistance significantly improved overall technical performance (OSATS 19.79±4.06 vs. 17.32±4.11; p=0.027). This gain was experience-dependent; AI significantly augmented trainee performance (19.20±3.76 vs. 16.60±3.78), narrowing the proficiency gap, while expert performance remained high and stable. 100% of participants identified the system as a useful training tool. However, subjective workload was significantly higher in the AI arm (SURG-TLX 26.42±9.56 vs. 22.26±7.81; p=0.014). Despite this, usability (SUS 75.13±14.31) and implementation feasibility, acceptability, and appropriateness scores were consistently high (means >4.4/5).

**Conclusions:** This study provides a stepwise process for real-time AI development using pituitary surgery as a high-stakes exemplar. The refined surgeon-centric AI system improves training and technical performance, particularly for trainees. Next steps involve first-in-human studies and further exploration of longer-term human factors such as over-reliance, cognitive overload mitigation and trust calibration.

**Summary Sentence:** This study establishes a stepwise pipeline for real-time AI development in high-stakes surgery, exploring whether surgeon-centric navigation assistance augments surgeon performance and training, and providing a foundation for clinical translation.

## Background

Pituitary tumours are among the most common brain tumours, often causing significant morbidity^1^. Transsphenoidal surgery is the primary treatment, but it is high-risk as it requires continuous orientation within a narrow operative corridor involving critical structures, including the carotid arteries and optic nerves. Thus, complications are common, for example, vascular injuries (up to 1–2%) or incomplete resections (up to 70% of cases)^2-4^. These complications can cause death, disabling stroke, or require radiotherapy due to residual tumour.

To reduce these risks, surgeons rely on adjuncts to help recognise and protect important anatomical structures. However current adjuncts, such as image-guidance and Doppler ultrasound, have limitations; for example, surgical workflow disruption, registration error and intra-operative tissue shifts adaptivity^5^. Computer vision AI offers the potential for real-time anatomical recognition, overlaying it directly on surgical video, potentially mitigating these limitations^5^.

Recently, several computer vision AI models capable of accurate structure segmentation from pituitary surgery videos, including recognition of surgical targets and the critical structures that must be protected, have been developed^6-12^. Computer-based simulation studies have demonstrated the potential for these models to augment surgeon performance via improved anatomical structure recognition, particularly for surgeons-in-training^13, 14^.

To safely translate these real-time AI models from the digital world to the real world, factors beyond model accuracy must be considered^15^. Firstly, the surgeon-AI interface requires careful design, specifically tailored to needs of users - expert surgeons and surgeons-in-training^15^. Recent evidence suggests that information balance (AI output relevance vs. cognitive overload) and informed decision integration via AI prediction confidence metrics are important for surgeons performing pituitary surgery^16, 17^. Additionally, prior to real world deployment, the IDEAL guidelines for systematic clinical translation of surgical technology emphasize the importance of stepwise evaluation, including robust preclinical evidence encompassing both clinical impact and human factors, to drive iterative technology development.

Therefore, in this two-phase study, we deployed real-time anatomy segmentation computer vision AI models on a high-fidelity simulated surgery and sought to: 1) iteratively tailor the system to surgeons’ needs, and 2) then evaluate its benefits for surgical navigation through a randomized control trial. We use endoscopic pituitary surgery as a high risk exemplar, with a pipeline that can be conceptually translated to other surgical procedures.

## Methods

### Overview

The study had two phases: 1) exploratory non-comparative physical simulation studies with the goal of iterative development of the AI model and device interface, and 2) a preclinical randomised crossover comparative trial of surgical performance with and without the version-fixed live AI assistant. The study was guided by the IDEAL guidelines and was approved by an institutional ethics committee (UCL 26117/002)^18, 19^.

### Setting

The mock operating theatre used was located at the National Hospital for Neurology and Neurosurgery (Figure 1). It was equipped with a full endoscopic pituitary surgery set, including drills and suction, and a previously validated high-fidelity pituitary surgery simulator (UpSurgeOn TNS Box) with replaceable disposable cartridges manufactured in-house^20^. These cartridges had the approach phase of the operation complete, such that the participants could proceed directly to the simulated tumour resection consisting of three steps: bone removal (sellotomy), dural opening (durotomy) and tumour resection. The morphology of the sella, internal carotids, optic nerves and sphenoid sinus were the same in each cartridge.

**Figure 1.**
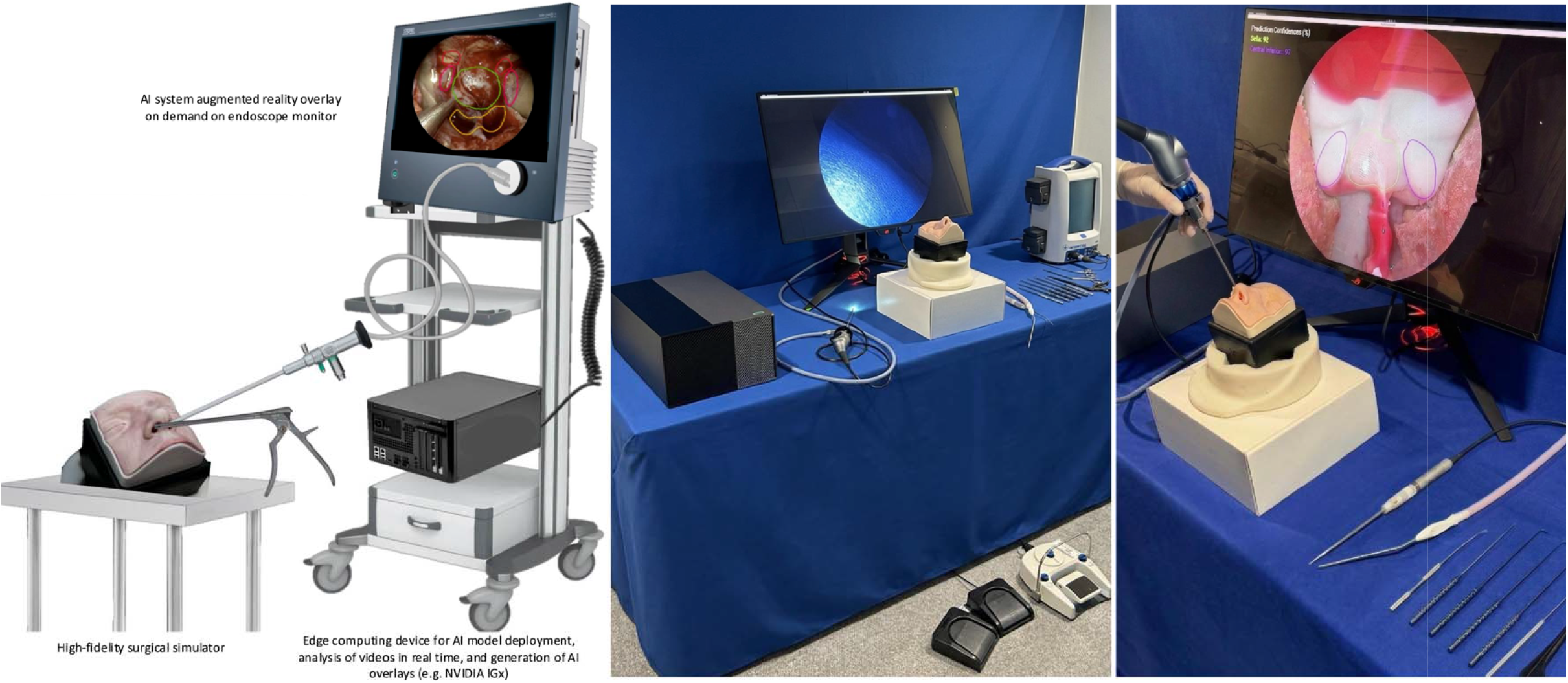
A: Overview of system configuration. B&C: Mock operating theatre set-up, with left panel showing surgical monitor, endoscope, high-fidelity simulator, surgical instrument set, edge computing device and foot pedals for controlling the AI overlays (left sided dual pedal) and drill (right sided pedal). The right panel shows a view from within the simulator of the start of each simulated surgery, with the AI overlay toggled on for the sella and carotid arteries.

Trainee (junior/mid-level residents) and expert-level (attendings/senior fellows) neurosurgeons were recruited via convenience sampling locally. There were no exclusion criteria.

The AI models were deployed on a high-performance edge computing device (NVIDIA AGX and IGX Orin), with its initial version based on the published ConsisTNet model, with additional training on simulator data^7, 8, 21^. The endoscope feed was passed directly into the edge computer, which displayed AI overlays (sella, carotid arteries, optic nerves and clival recess) on the primary surgical monitor and was controlled with a foot pedal (Figure 1; Video 1). The AI-surgeon interface had two key features, derived from precursor surgeon survey studies: 1) the structures were outlines (balancing information pertinence vs. overload) and 2) surgeons were in full control of the activation of the AI display via a foot pedal (no automation, again, to control information flow)^16, 17^.

### Phase 1: Exploratory Simulation Study

The aim of this phase was to obtain iterative feedback from a small group of surgeons to drive iterative improvements in the prototype AI model and AI-surgeon interface. The end point was achieving a fixed prototype which could be assessed via comparative trial, with no remaining performance-limiting issues highlighted via user feedback.

Surgeons were asked to perform a simulated pituitary surgery with access to the AI assistance (Figure 2). During the procedure, surgeons were encouraged to use the AI assistant and give feedback via a Think Aloud protocol, with notes taken by the study team. At specific checkpoints (pre-sellotomy, pre-durotomy, and pre-tumour resection), surgeons were asked to comment on the anatomical accuracy of the AI overlays.

**Figure 2.**
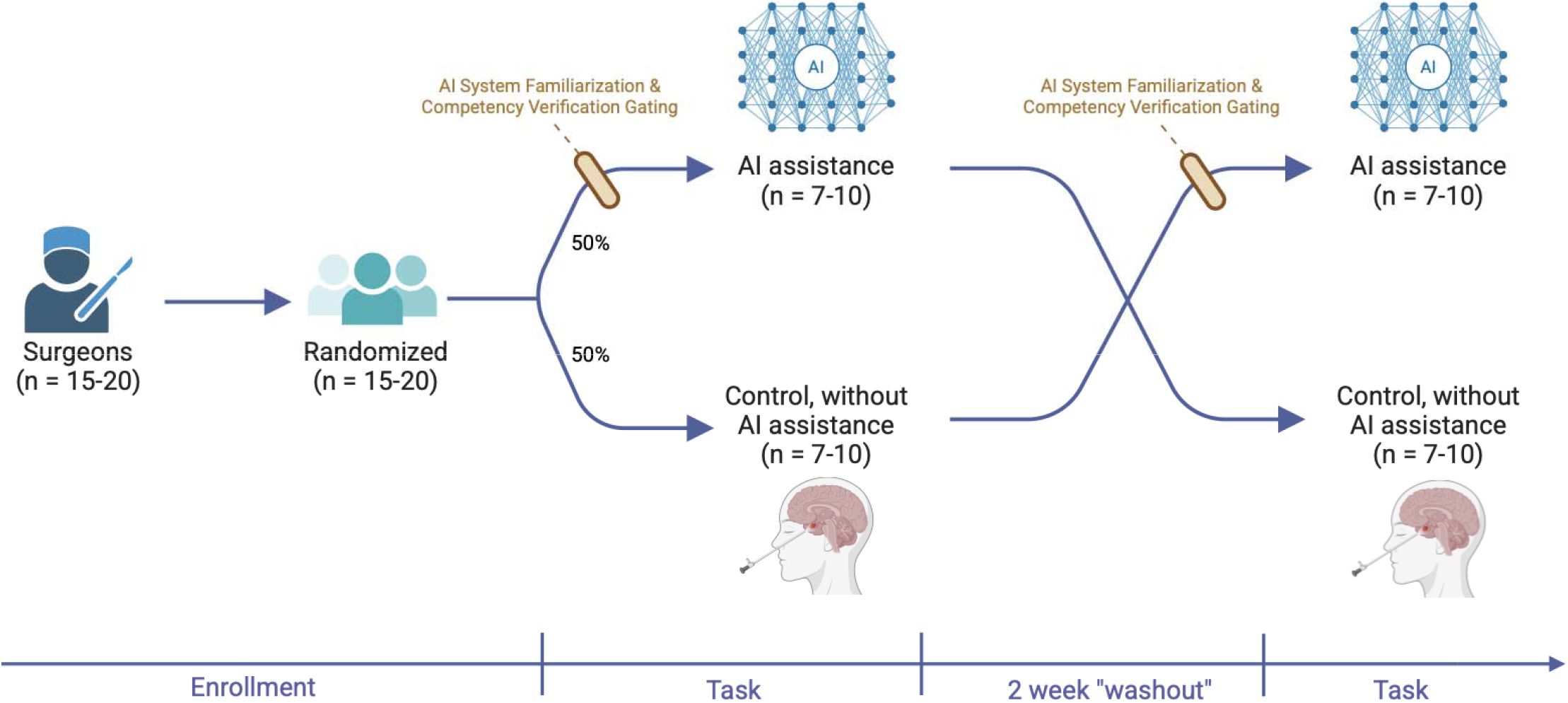
Overview of cross-over trial design. Randomization was done via 50:50 permuted block randomisation. Each trial began with a demo by a study team member of the surgical equipment, surgical simulator, +/-AI system. The AI system demo included explanation of the AI model and the envisioned point of deployment as an anatomy verification tool; AI toggling on and off; cycling through various display options; toggling on confidence metrics; and guided interpretation of the AI model output. This was followed by a “familiarisation” assessment which involved asking the users to demo all taught features. Finally, a “competency” assessment was performed to assess the interpretation of the AI model output, which included turning on all AI features (i.e. toggling on all structures with associated confidence metrics) and commenting on whether the displayed anatomical segmentation aligned with the true anatomy. Responses classified into pass (appropriately agrees with accurate segmentation and appropriately disagrees with inaccurate segmentation) and fail (inappropriately agrees or disagrees with accurate segmentation), and appropriateness with judged on clinical anatomical reasoning (for example, “I disagree with the segmentation of the right carotid as I see it does not include an expected area inferior to the optico-carotid recess”). The competency assessment was performed with alternating accurate and deliberately less accurate AI models, and three consecutive passes were required to proceed into the AI-assisted arm.

Peri-procedure questionnaires captured demographics (age, sex, level of surgical training, pituitary surgery experience), baseline AI literacy (via Meta AI literacy scale; MAILS), system usability (via System Usability Scale; SUS), trust in the system (via Short Trust in Automation Scale; STIAS), procedure workload (Surgery Task Load Index; SURG-TLX), and qualitative feedback for improvements to system usability, trust and workload (Appendix 1). Basic descriptive statistics were used for continuous and categorical data. Qualitative data from the Think Aloud protocol notes and questionnaire feedback were thematically categorised into feedback about: the AI model, the AI-surgeon interface and other issues.

### Phase 2: Randomised Cross-over Comparative Trial

The aim of this phase was to assess an updated prototype after the phase 1 study on surgical safety, performance and training. Therefore, a randomised cross-over trial design was chosen, where surgeons perform a simulated endoscopic pituitary tumour resection (sellotomy, durotomy and tumour resection), with two arms (Figure 2) - one with AI system assistance and one without. Entry to the AI-assisted arm was gated by a demo and a system familiarization assessment to ensure users understood the system and how to effectively use it (Figure 2, Appendix 2)^22, 23^. A minimum 2-week washout between the trial arms was implemented to mitigate surgical learning curve and AI-assistance carryover effects. The prototype was fixed (hardware, AI model and user interface) for the duration of the trial. An *apriori* sample size estimation suggested recruitment of 15-20 surgeons^24^.

The primary outcome was surgical performance and safety, measured using the validated Objective Structured Assessment of Technical Skills (OSATS) scale, adapted for single surgeon pituitary surgery and retrospective blinded video by a single attending neurosurgeon to maintain internal consistency (HJM)^20, 25^. The same quantitative data collected in Phase 1 was collected in Phase 2 (demographics, MAILS, SUS, SURG-TLX, STIAS). In addition, educational yield (via structured questionnaire; Appendix A), feasibility (Feasibility of Intervention Measure; FIM), Acceptability (Acceptability of Intervention Measure; AIM) and Appropriateness (Intervention Appropriateness Measure; IAM)^26-29^. Furthermore, exploratory outcomes were assessed at each checkpoint (pre-sellotomy and pre-durotomy): alignment with the AI segmentation (“How much do you agree that the outlines provided by the AI align with the correct anatomy? Strongly/partly/disagree, and explain”), influence of surgical decision making confidence (“After using the tool, has it changed your confidence in your surgical decision making? Increase/decrease/no change, and explain”)^30^.

Basic descriptive statistics were used for continuous and categorical data from questionnaires, checkpoint questions and video review. Comparative statistics were performed to compare performance and safety (OSATS) and workload (SURG-TLX) in both arms, via linear mixed-effects modelling, adjusting for the trial sequence (arm order) as a fixed effect and participant as a random effect. This was done overall, and then per experience level-sub group.

## Results

### Phase 1: Exploratory Simulation Study

Phase 1 consisted of two exploratory sessions, with each session followed by a period of system iteration - with the end point a stable system deemed ready for comparative analysis. Four surgeons (3 trainees, 1 attending) participated in the first test and four surgeons (4 trainees) participated in the second test. One participant was in both trials. Participant characteristics, quantitative survey scores and qualitative feedback summary (categorised into hardware, AI model, and user interface changes) are shown in Tables 1 and 2.

**Table 1.**
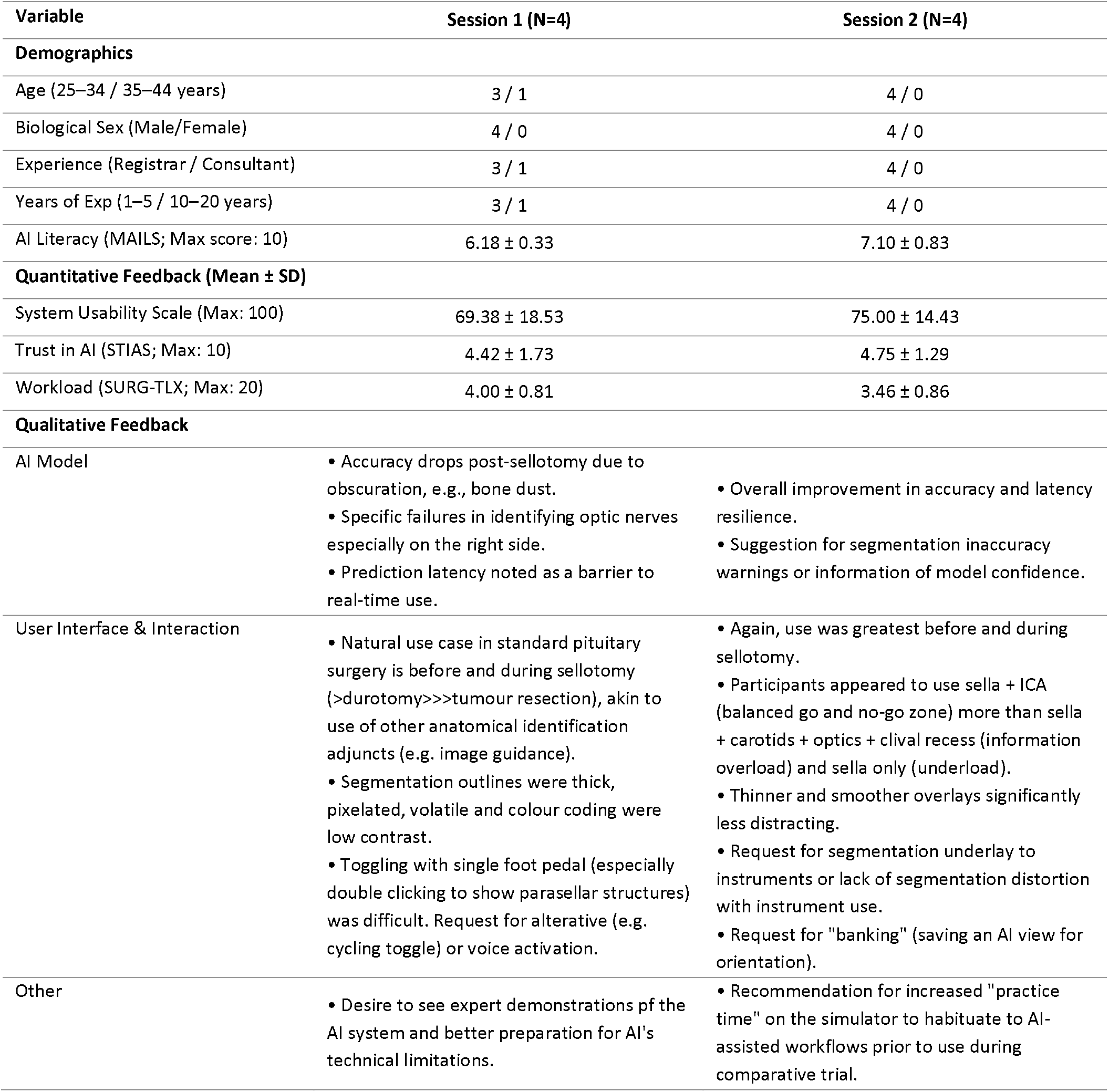
Over of Phase 1 Study Results.

| Variable | Session 1 (N=4) | Session 2 (N=4) |
| --- | --- | --- |
| **Demographics** |  |  |
| Age (25–34 / 35–44 years) | 3 / 1 | 4 / 0 |
| Biological Sex (Male/Female) | 4 / 0 | 4 / 0 |
| Experience (Registrar / Consultant) | 3 / 1 | 4 / 0 |
| Years of Exp (1–5 / 10–20 years) | 3 / 1 | 4 / 0 |
| AI Literacy (MAILS; Max score: 10) | 6.18 ± 0.33 | 7.10 ± 0.83 |
| **Quantitative Feedback (Mean ± SD)** |  |  |
| System Usability Scale (Max: 100) | 69.38 ± 18.53 | 75.00 ± 14.43 |
| Trust in AI (STIAS; Max: 10) | 4.42 ± 1.73 | 4.75 ± 1.29 |
| Workload (SURG-TLX; Max: 20) | 4.00 ± 0.81 | 3.46 ± 0.86 |
| **Qualitative Feedback** |  |  |
| AI Model | • Accuracy drops post-sellotomy due to obscuration, e.g., bone dust.<br>• Specific failures in identifying optic nerves especially on the right side.<br>• Prediction latency noted as a barrier to real-time use. | • Overall improvement in accuracy and latency resilience.<br>• Suggestion for segmentation inaccuracy warnings or information of model confidence. |
| User Interface & Interaction | • Natural use case in standard pituitary surgery is before and during sellotomy (>durotomy>>>tumour resection), akin to use of other anatomical identification adjuncts (e.g. image guidance).<br>• Segmentation outlines were thick, pixelated, volatile and colour coding were low contrast.<br>• Toggling with single foot pedal (especially double clicking to show parasellar structures) was difficult. Request for alternative (e.g. cycling toggle) or voice activation. | • Again, use was greatest before and during sellotomy.<br>• Participants appeared to use sella + ICA (balanced go and no-go zone) more than sella + carotids + optics + clival recess (information overload) and sella only (underload).<br>• Thinner and smoother overlays significantly less distracting.<br>• Request for segmentation underlay to instruments or lack of segmentation distortion with instrument use.<br>• Request for "banking" (saving an AI view for orientation). |
| Other | • Desire to see expert demonstrations of the AI system and better preparation for AI's technical limitations. | • Recommendation for increased "practice time" on the simulator to habituate to AI-assisted workflows prior to use during comparative trial. |

**Table 2.** Participant Characteristics for Cross-over Comparative Trial.

| Characteristic | Total Cohort (N=19) |
| --- | --- |
| **Age (years)** |  |
| 25–34 | 13 (68.4%) |
| 35–44 | 5 (26.3%) |
| 45–54 | 1 (5.3%) |
| **Biological Sex** |  |
| Male | 17 (89.5%) |
| Female | 2 (10.5%) |
| **Surgical Experience Level** |  |
| Junior Resident (ST1-3) | 8 (42.1%) |
| Senior Resident/Registrar (ST3+) | 8 (42.1%) |
| Expert (Fellow/Consultant) | 3 (15.8%) |
| **Expert Status (per Protocol)** |  |
| Trainee | 15 (78.9%) |
| Expert | 4 (21.1%) |
| **Pituitary Surgery Experience** |  |
| < 1 year | 11 (57.9%) |
| 1–5 years | 7 (36.8%) |
| 5–10 years | 1 (5.3%) |
| **Baseline AI Literacy (Mean MAILS Score; Total: 10)** | 6.43±0.72 |

Notably, although the usability and trust appeared to improve and workload appeared to decrease after changes made between Session 1 and 2, the small heterogeneous sample size and related confounders preclude comparative analysis. The focus is instead on actionable insights gained from observation, think aloud protocol and qualitative survey data, which is the driver for iterative system improvements (Table 2; Appendix 3).

Using the Session 1 feedback, several hardware, AI model and user interface improvements were implemented. To improve latency, a more powerful computing device was chosen (NVIDIA IGx) and the model architecture was changed (to an DINOv3-based vision transformer with a Tensor RT optimization)^31^. To address AI model accuracy, particularly when structures were partially obscured or viewed from the left nostril, further binasal video data was annotated, including frames with bone dust, wash, etc. to improve model performance in these surgical scenes. Regarding interface changes, these included thinner structure outlines, smoothing functions to reduce volatility and better outline colour contrast. The foot pedal triggering strategy was changed from click number matching (e.g., single click = sella, double click = carotids, etc.) to incremental cycling (single click = sella, further single click = sella + carotids, then sella + carotids + optic nerves + clival recess; Video 1).

Using Session 2 feedback, further refinements were made. This included further data annotation, particularly in obscured surgical scenes, to further improve AI model performance. AI model confidence for each structure was introduced as a per-structure score (0–100), calculated by aggregating the model’s prediction probabilities (post-Softmax) across all pixels belonging to each segmented structure. The foot pedal was changed to a 2 panel foot pedal such that the second panel was used to toggle the confidence metrics on/off. Requests for structured demonstration and more familiarisation with the system prior to use was implemented into the comparative trial (see Methods). The pre-tumour resection checkpoint was removed given user feedback that the tool was unlikely to be helpful at this stage. The request for overlays that remain unaffected by occlusions from surgical instruments was recognized as clinically important, but is not feasible within the current real-time semantic segmentation framework and available annotations. Enabling this capability would likely require techniques such as amodal segmentation, together with substantially more annotated data to facilitate stronger contextual reasoning. However, these additions would likely increase computational complexity and make it more challenging to satisfy intraoperative latency constraints.

### Phase 2: Randomised Cross-over Comparative Trial

Twenty surgeons were recruited to the trial, 16 trainees and 4 experts, with their characteristics shown in Table 2. One participant (trainee) was excluded from the final analysis due to being a statistical outlier on OSATS scoring (Z-score>2.5) and encountering a technical video capture failure during their initial trial arm, which introduced significant learning-curve bias upon repeat. Consequently, 19 participants were included in the comparative analysis. All participants passed the familiarisation and competency assessment when in the AI arm.

#### AI impact on performance

The use of real-time AI assistance significantly improved performance, as measured by OSATS (19.79±4.06 with AI vs. 17.32±4.11 without; p=0.027 when adjusted for order; Figure 3 & Appendix 5). No significant effect was found for arm order (p=0.593), suggesting the success of the 2-week washout period. Subgroup analysis demonstrated that the performance gain was most pronounced among trainees (19.2±3.76 with AI vs. 16.6±3.78 without). Experts maintained consistently high scores regardless of assistance (22.0±4.97 with AI vs. 20.0±4.76 without). One expert declined from OSATS score 27 to 20 (still greater than the mean performance of trainees with AI). The mean difference in OSATS between arms was +2.47 overall, +2.6 in trainees, and +2 in adults.

**Figure 3.**
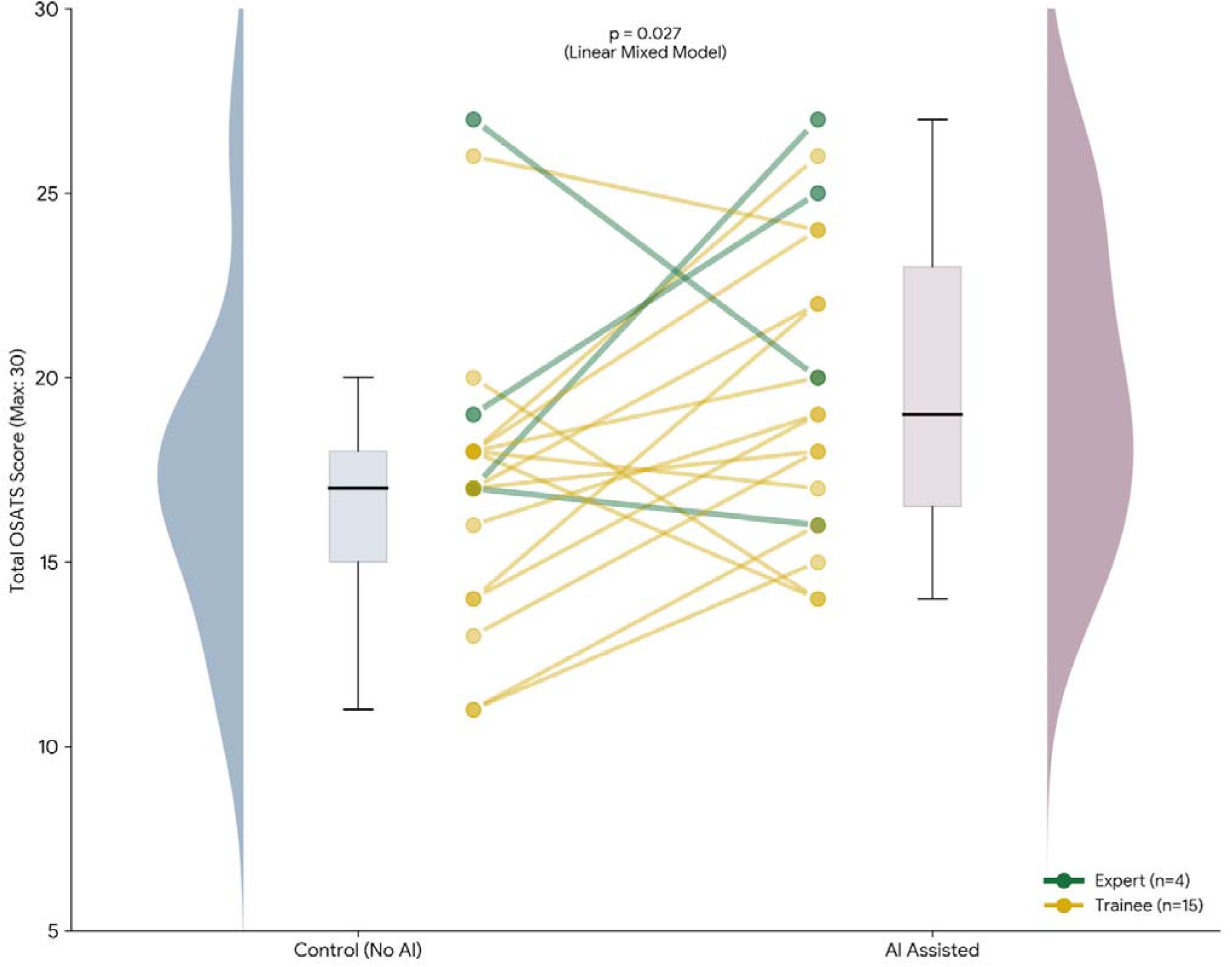
Comparative Analysis of Surgical Performance (OSATS). Raincloud plot displays the density distribution and box-and-whisker display summary statistics for the Control arm (Deep Blue) and AI Assisted arm (Deep Plum). Individual participant performance shifts are shown via connected dots, with line thickness indicating surgical role (Thick Blue = Trainee, n=15; Thick Red = Expert, n=4). Statistical significance was determined using a linear mixed-effects model adjusted for trial order (p=0.027), with the p-value label positioned equidistant between the paired columns.

#### AI impact on training

All participants (n=19) identified the AI assistant as a useful training tool that reinforced clinical knowledge by verifying anatomical landmarks and highlighted skill gaps in action (Appendix 4).

#### AI-surgeon interaction

Surgeon perception of the system fluctuated across pre-sellotomy and pre-durotomy, with trust remaining despite reducing AI accuracy agreement and decision confidence impact. At pre-sellotomy (n=19), 68.4% strongly agreed and 31.6% partly agreed with AI accuracy; 68.4% reported increased and 31.6% unchanged decision confidence; and mean trust in AI (STIAS, max 6) was 5.09±0.88. At pre-durotomy (n=18), 52.6% strongly agreed and 42.1% partly agreed with AI accuracy; 42.1% reported increased and 52.6% unchanged decision confidence, with a mean trust of 5.37±0.89. There was no instance of total AI accuracy disagreement or decreased decision confidence at either timepoint. One participant was excluded at pre-durotomy due to inadvertent early durotomy during sellotomy.

However, subjective workload was significantly higher in the AI-assisted arm (26.42±9.56) compared to the control arm (22.26±7.81; p=0.014). Despite this, the system achieved a mean System Usability Scale (SUS) score of 75.13±14.58, meeting the criteria for “Good” usability.

#### AI implementation metrics

Implementation feasibility and acceptability scores remained consistently high across validated scales, with a FIM of 4.53±0.45 (max 5), AIM of 4.43±0.67 (max 5) and IAM of 4.45 ±0.52 (max 5).

## Discussion

### Principal Findings

This study demonstrates the successful translation of a computer vision model into a real-time, surgeon-controlled prototype following systematic and iterative refinements in AI architecture, interface, and hardware. Our primary finding is the “leveller” effect of AI assistance - significantly improved technical performance (OSATS) for trainees (19.2±3.76 vs. 16.6±3.78; p=0.027), narrowing the gap between trainees and experts without compromising the performance of senior surgeons, who maintained a high baseline. When compared with real-world in-vivo OSATS benchmarks for the sellar phase (mean: 20.9, SD: 2.1), our control expert cohort demonstrated comparable performance (mean: 20, SD: 4.76), whilst our trainee cohort moved from a mean performance two SDs below the real-world mean, to within one SD^32^.

However, this gain was accompanied by an increase in subjective workload, despite high acceptability, usability, feasibility and appropriateness scores. Further study is needed to see whether this “cognitive tax” is sustained with repeated use, and if so, mitigation strategies should be developed^33^. Furthermore, system reliability proved phase-dependent, with accuracy agreement and decision confidence declining post-sellotomy as environmental “noise”, such as bone dust and irrigation, obscured the surgical field. This accuracy decline resulted in a lack of impact on surgical decision-making confidence but the clinical impact of this was not borne in overall OSATS scoring – probably because the clinical yield of the anatomical navigation assistance was lower post-sellotomy anyway. The sustained, and sometimes increased, surgeon trust in the AI system across these checkpoints despite this accuracy reduction is likely due to the presentation of AI prediction confidence metrics, which reduced with poor accuracy predictions, and established a transparent relationship with the surgeon user^17, 34^.

### Findings in the Context of the Literature

The role of AI as a potential “leveller” in surgical decision support between expertise groups but a “discriminator” between human alone and human + AI groups has been demonstrated in previous digital simulation studies in our group^13, 14^. The relative lack of improvement in performance in the expert group, previously thought to be related to sub-optimal human factors or design issues^15^, was reproduced here despite systematic iterative user-centred development of this system^16, 17^. This probably represents limitations of applied surgical AI, which does not yet have the experience (biased and/or small training datasets) or multimodal data abilities that an expert human surgeon has, and therefore experts have less to gain from this system in its current form^35-37^. Despite that, the educational yield, particularly for the experts to teach their trainees, was acknowledged, with this sentiment reflected in the wider literature as the first translational benefit for many surgical AI systems^37-40^.

In other surgical fields, real-time computer vision based AI systems have shown promise, despite only making up 12% (n=13) of surgical computer vision studies, with this number set to grow as this novel technology is translated into clinical practice^41^. Most surgical computer vision studies investigate the use of anatomy recognition models (as opposed to workflow recognition, instrument recognition or other tasks) and all were in general (mostly laparoscopic) surgery domains^41^. Only a handful of studies have reached prospective clinical stages^41^. For example, an early clinical study in laparoscopic cholecystectomy, where AI segmentation of anatomical structures to highlight the “critical view of safety”, demonstrated technical feasibility and early promise as a real-time intra-operative training and decision support tool^42-46^. Similar promise for clinical decision support and training was demonstrated in nerve recognition during laparoscopic colorectal surgery^47^. However, no such studies were performed in neurosurgery, which is arguably higher stakes regarding complications related to surgical adverse events^41^. Translation in such high stakes environments requires a systematic iterative approach, which goes beyond AI model performance, clinical impact and feasibility, and includes deliberate user-centred design, human factors analysis and robust pre-clinical simulation studies prior to clinical translation. This is reflected in the principles of the IDEAL and DECIDE-AI frameworks, and thus this present study demonstrates an exemplar pipeline for preclinical development and evaluation, particularly for higher stakes surgeries, but also for other surgeries in which there is a paucity of this approach^18, 19, 48^. Several human factors require longer term studies, including the development of appropriate trust, risk of deskilling over time if surgeons are over reliant, and the mitigation of cognitive overload as discussed previously.

### Strengths and Limitations

The major strength of this study is its systematic approach to user-centred AI platform design, with a randomized crossover validation on a high-fidelity simulator. However, several limitations remain. The expert subgroup (n=4) was small, limiting the power of subgroup statistics. Additionally, while the simulator is high-fidelity, it cannot fully replicate dynamic challenges like major haemorrhage or the reality of patient-specific anatomical variability, which may further degrade AI model performance. Finally, this study was based at a single centre, so its external validity is not certain.

## Conclusion

This study provides a stepwise pipeline for real time AI system development, using pituitary surgery as a high-stakes surgical exemplar. We have demonstrated that a surgeon-centric AI anatomical navigation assistance system can improve surgical education and operative performance, particularly in trainees. Having met the requirements for IDEAL Stage 0, including high usability and feasibility metrics, this system is prepared for IDEAL Stage 1 first-in-human clinical studies. Future iterations must prioritize reducing cognitive load and monitoring for over-reliance in challenging cases with lower AI accuracy or confidence, and over time with repetitive AI use.

## Data Availability

Available upon reasonable request

## Figures, Tables and Video

Video 1: https://liveuclac-my.sharepoint.com/:v:/g/personal/rmapdk0_ucl_ac_uk/IQCobAWTU6vHSr9Zupygr5EiAarB8rvBuRfewq3f7ZoKwMc?e=OGfvOo

## Appendices

## APPENDIX 1 User surveys

## APPENDIX 2 Study member survey for comparative study (including familiarisation assessment)

## APPENDIX 3 Phase 1 dataset, including qualitative data

## APPENDIX 4 Phase 2 dataset, including qualitative data

## APPENDIX 5 OSATS by Subdomains. Comparative analysis via mixed effects linear model, adjusting for the trial sequence (arm order) as a fixed effect and participant as a random effect. *Statistically significant (p<0.05).

## Notes

**Conflicts of Interest** HJM is employed by and hold shares in Panda Surgical. DS holds shares in Panda Surgical, Odin Vision, and is employed by TouchSurgery, Medtronic.

**Funding** This work was funded by the EPSRC (EP/Y01958X/1 and EP/W00805X/1, UKRI145). It was supported more generally by the UCL Hawkes Institute, formerly the Wellcome/EPSRC Centre for Interventional and Surgical Sciences (WEISS) (203145/Z/16/Z). DZK is supported by an NIHR Doctoral Fellowship and Google PhD Fellowship. HJM is supported by the NIHR Biomedical Research Centre at University College London. DS is supported by the Department of Science, Innovation and Technology (DSIT), and the Royal Academy of Engineering under the Chair in Emerging Technologies programme.

### Competing Interest Statement

HJM is employed by and hold shares in Panda Surgical. DS holds shares in Panda Surgical, Odin Vision, and is employed by TouchSurgery, Medtronic.

### Author Declarations

Approved by an institutional ethics committee at University College London (UCL 26117/002)

